# Developing an Evaluation Framework for Infectious Disease Modelling-to-Policy Pathways: A Qualitative Study Across Five Continents

**DOI:** 10.64898/2026.07.22.26358658

**Authors:** K Röbl, EN Iftekhar, K Oliver, HT Fischer, S Funk, J Fitzner, J Hanefeld

## Abstract

Infectious disease modelling has become an increasingly prominent tool in public health decision-making, with its use accelerating markedly during the COVID-19 pandemic. This growth calls for an understanding of how modelling evidence is received, interpreted, and used – or not used – by decision-makers. There is a recognised need for evaluations of modelling-to-policy systems to understand how to best integrate modelling evidence into decision-making. So far, no comprehensive evaluation framework exists that maps modelling-to-policy pathways. This study addresses that gap by developing a theory of change for modelling-to-policy systems that could serve as a foundation for future evaluations. A qualitative study design was employed, comprising semi-structured interviews with 35 modellers, knowledge brokers, and decision-makers across five continents and diverse institutional settings, spanning high-income and low- and middle-income countries, as well as national and international modelling-to-policy contexts. Thematic analysis was combined with a backward-mapping-informed approach to develop a multi-level evaluative framework. The protocol for this study has been published on March 20, 2025, at OSF (https://doi.org/10.17605/OSF.IO/J9QXV). Participants diverged in their conceptualisations of successful modelling evidence use – ranging from instrumental use to accurate understanding and consideration of modelling outputs – yet converged on shared risks: decisions informed by inadequately specified models or by evidence that is misinterpreted due to communication failures. The resulting three-level framework identifies factors directly influencing modelling evidence use across three domains (policy relevance, model quality, and communication and interaction), traces these to enabling conditions, and maps them to systemic enablers – including local and embedded modelling capacity, data infrastructure, knowledge brokering capacity, formal knowledge translation structures, established networks, and funding. The proposed framework represents a first theory of change for modelling-to-policy systems. While it requires further testing and application across diverse decision-making contexts, it offers a structured basis for evaluating existing systems and informing the design of new ones.

## INTRODUCTION

Infectious disease modelling is a mathematical approach to representing disease dynamics in ways that go beyond descriptive statistics or regression-based analyses. Where conventional epidemiological methods characterise associations within observed data, mathematical models can represent the dynamic, mechanistic processes underlying disease transmission – enabling the estimation of outcomes that cannot be directly observed, such as the projected impact of a planned intervention or the comparison of consequences of alternative policy scenarios. (James et al., 2021) As such, modelling can inform decision-making in public health with regards to infectious diseases: it supports evidence synthesis, outbreak forecasting, and the formal comparison of decision alternatives, ranging from outbreak control strategies to the design and roll-out of vaccination programmes. (Heesterbeek et al., 2015; Lancet Commission on Strengthening the Use of Epidemiological Modelling of Emerging and Pandemic Infectious Diseases, 2024)

The use of infectious disease models to inform public health decision-making has grown substantially over recent decades, a trend accelerated by the COVID-19 pandemic. Across many countries, policymakers drew on modelling outputs to guide their responses – bringing the discipline into unprecedented public and political visibility. (James et al., 2021; Lancet Commission on Strengthening the Use of Epidemiological Modelling of Emerging and Pandemic Infectious Diseases, 2024) This growing influence makes it increasingly important to understand how modelling evidence is received, interpreted, and used – or not used – by decision-makers. There is a need for evaluations of modelling-to-policy systems to understand how to maximize the utility of modelling evidence for decision-making; yet, to date, no such evaluations exist. (Rao et al., 2025)

At the same time, this increased prominence has exposed challenges around the use of infectious disease modelling evidence. Depending on the underlying data quality, modelling outputs can be highly uncertain, and for novel pathogens many parameters remain poorly understood. (Heneghan & Jefferson, 2022; Holmdahl & Buckee, 2020) Often multiple defensible approaches to model construction exist, consequently results may diverge, complicating the interpretation of results. (James et al., 2021) Decisions about model structure are themselves subjective, yet structural uncertainty is rarely communicated to decision-makers – reported confidence intervals typically capture only one dimension of uncertainty, risking a false sense of precision, (Holmdahl & Buckee, 2020; James et al., 2021) while the absence of standardised reporting guidelines leaves these risks largely unmitigated. (Heneghan & Jefferson, 2022) These challenges underline the importance of understanding how risks to effective modelling evidence use can be prevented.

The contribution of research evidence to decision-making and its barriers and enablers have been studied across a range of evidence types, and approaches exist to evaluate and support these processes. (Nutley et al., 2007; Oliver et al., 2014; Salter & Kothari, 2014; Weiss, 1998) A substantial body of work on evidence-to-decision frameworks has sought to conceptualise and improve the pathways through which research evidence informs policy, generally pursuing one or more of the following: describing how evidence moves from production to use; explaining how and why it is or is not used; evaluating evidence use in a given context; or improving the process through practical guidance. (Kitson et al., 2008; Parkhurst et al., 2021; Rinnert, 2017; Sempé et al., 2025) This literature commonly identifies three main actor groups in evidence-to-policy systems: evidence producers (such as researchers); knowledge brokers or intermediaries (who link researchers and decision-makers); and knowledge users (such as civil servants or policymakers). (Knight & Lyall, 2013; Sempé et al., 2025)

There have been first attempts to develop such frameworks specifically for modelling evidence, setting out pathways for how scenario modelling can support decision-making. (Androutsopoulou & Charalabidis, 2018) However, this work is not empirically grounded and does not identify the influential factors or necessary preconditions that shape whether modelling evidence is actually used. Such factors are needed to evaluate modelling-to-policy pathways and to guide the structural design of modelling-to-policy systems. (Rao et al., 2025) A growing body of literature has begun to examine facilitators and barriers to infectious disease modelling evidence use in public health decision-making, (Dhungana et al., 2025; Hadley et al., 2025a; Owek et al., 2024; Pepin et al., 2025; Rao et al., 2025) but this work is largely confined to emergency and outbreak contexts. Comparatively little is known about how modelling informs more routine decision-making – for example, on vaccination schedules or programmes. Additionally, while studies began to identify relevant facilitators and barriers, they do not provide the logical chains needed to understand how structural inputs lead to use and utility of modelling. This is a gap that the implementation research and programme evaluation literature has long addressed through logic models and theories of change. These tools describe the hypothesized causal pathways through which inputs and activities are expected to produce outcomes, enabling evaluators not only to assess whether an outcome has been achieved but also to understand why it has or has not, and what preconditions are required. (Funnell & Rogers, 2011; McLaughlin & Jordan, 2015) Theories of change have been applied in the evidence-use literature to evaluate knowledge translation strategies. (Kreindler, 2018; Minogue et al., 2021; Sell et al., 2024) However, to our knowledge, no such theory of change or logic model has been developed for infectious disease modelling-to-policy pathways.

A further gap concerns the structural dimension of modelling-to-policy systems. Hadley et al. have produced valuable insights here with their typology mapping configurations of how modelling teams and decision-making bodies relate to one another, (Hadley et al., 2025a) the number of and collaboration between modelling teams that inform national decision-makers, and their level of government interaction ranging from medium to embedded, i.e. modellers being employed by decision-making bodies. Recent work has also begun to identify structural factors perceived as enabling modelling evidence use in low- and middle-income country contexts specifically. (Oliwa et al., 2025) However, this work is limited to pandemic settings and does not extend to routine decision-making, and while it identifies relevant structural enablers, it does not provide a comprehensive account of the pathways through which these factors lead to perceived successful modelling evidence use. Moreover, Hadley et al.’s, their typology does not explicitly account for settings in which modellers operate in geographically distinct locations from the decision-makers they support – a configuration that is common in low- and middle-income country contexts, where international modelling teams frequently inform national decision-making. (Aguas et al., 2020) Moreover, the typology does not express a preference for any particular system configuration (Hadley et al., 2025a), leaving open the question of how structural factors shape modelling-to-policy outcomes and what structural interventions might improve them.

Finally, the existing literature on facilitators and barriers to modelling evidence use is predominantly produced by modellers, and thus largely reflects the perspectives of evidence producers rather than users. This is a meaningful limitation: the broader evidence-use literature suggests that producers and users often hold divergent views on what constitutes successful evidence use, with producers tending to underestimate the complexity of policymaking processes. (Cairney, 2016; Greer, 2022) Understanding how decision-makers perceive and engage with modelling evidence is therefore essential, yet remains underexplored.

In summary, while a growing body of literature examines facilitators and barriers to modelling evidence use in public health, no comprehensive theoretical framework yet exists that maps modelling-to-policy pathways starting from structural factors, spans both emergency and routine decision-making contexts, and integrates the perspectives of decision-makers alongside those of modellers. Given the increased use of infectious disease modelling in public health decision-making, there is a need to understand where and how modelling evidence utility can be optimised, and which structural interventions are most likely to improve system design.

This study addresses this gap by developing a theory of change for modelling-to-policy systems, drawing on qualitative interviews with modellers, knowledge brokers, and decision-makers across diverse public health contexts. We identify key enabling factors of perceived successful modelling evidence use, map these across three levels and propose causal mechanisms linking factors and levels. The resulting framework is intended to serve as a foundation for future evaluations of – and practical guidance for – modelling-to-policy systems.

Specifically, this study addresses the following research questions:

- What are modellers’, knowledge brokers’, and decision-makers’ perceptions of successful modelling evidence use?
- How can risks to the effective use of modelling evidence be prevented?
- What structural factors of modelling-to-policy systems facilitate modelling evidence use, and through what pathways and mechanisms?
- How do these pathways differ between outbreak and routine contexts, and national and international modelling-to-policy systems?

## METHODS

This study applied the Standards for Reporting Qualitative Research guidelines and adheres to the Consolidated Criteria for Reporting Qualitative Research checklist where applicable. (O’Brien et al., 2014; Tong et al., 2007). The protocol for this study has been published on March 20, 2025, at OSF (https://doi.org/10.17605/OSF.IO/J9QXV).

### Ethics statement

This research has been advised by the Ethics Committee of the Berlin Medical Association (Ärztekammer Berlin, process nr. ETH-SB-25-015-1) and approved by the Research Ethics Review Committee (ERC) of the World Health Organization (protocol ID. ERC.0004269). All participants received information about the study purpose and procedure, data protection and proposed outputs and provided written informed consent prior to participation.

### Study design

We employed a qualitative research design to gain an in-depth understanding of modelling-to-decision-making pathways. We conducted semi-structured interviews with key informants that are engaged or have been engaged in the past in modelling-to-policy processes, such as modellers (knowledge producers), intermediaries (knowledge brokers), and public health decision-makers (knowledge users). We defined

- **knowledge producers** as modellers who are producing or have produced mathematical modelling outputs in the past with the purpose of informing public health decision-making,
- **knowledge brokers** as persons who have been involved in communication between modellers and decision-makers, or the translation and adaptation of policy questions and modelling outputs, and
- **knowledge users** as persons being involved in public health decision-making on a technical or political level.

Participants could fulfil the definitions of more than one of these roles.

### Data collection instruments

The interview topic guides were developed based on two complementary inputs: the identification of knowledge gaps in factors influencing modelling-to-policy pathways through a literature search, and the outputs of an expert workshop convened by the WHO Hub for Pandemic and Epidemic Intelligence in October 2024 (World Health Organization, 2025), which brought together specialists to discuss key attributes for evaluating these pathways. (Rao et al., 2025) The topic guides are structured around modelling-policy interaction, the modelling process, modelling evidence use in decision-making and equity considerations as well as future outlooks and can be found in the supplementary files. The topic guides were not pilot tested before actual use in the study but refined through several feedback rounds within the study team.

### Participant recruitment and data collection

Participants were recruited through a) purposive sampling using the professional networks of the authors and the WHO Hub for Pandemic and Epidemic Intelligence to identify and approach potential participants, and b) snowball sampling asking participants to extend the study invitation to suitable and interested persons within their professional networks. (Biernacki & Waldorf, 1981; Palinkas et al., 2015) Purposive sampling was guided by an explicit aim for diversity – spanning different geographical backgrounds and genders, national and international modelling-to-decision-making systems, academic and government-employed modellers, and decision-makers across governmental and non-governmental organisations at both national and international levels. Contact was made via email in most cases and in few cases face-to-face.

Sampling continued until data saturation was reached. We defined data saturation as having reached both sample diversity – in terms of geographical background, national versus international settings, and gender – and thematic saturation, i.e. no new themes emerged during analysis.

Interviews were conducted in-person in a place where the interviewee felt comfortable speaking or online via the video-conferencing software Cisco Webex. The interviews were conducted without any non-participants present. In four instances, interviews were conducted jointly with two participants from the same institution, with three pairs being in a supervisor-subordinate relationship. While this format offered the advantage of capturing both aligned and diverging perspectives on shared experiences, it may also have introduced a degree of social desirability bias, potentially limiting participants’ willingness to express critical views. The interviews were scheduled for 45 minutes and their actual duration ranged from 35 to 58 minutes. Interviews were conducted in German, English or French. Almost all interviews were conducted jointly by KR and ENI. Exceptions included seven interviews conducted by KR alone, and one two-participant interview conducted by ENI alone.

No repeat interviews were carried out. All interviews were recorded for subsequent transcription. The interviews conducted online were video and audio recorded, the in-person interviews were only audio recorded. The transcripts were returned to participants for comments and corrections.

### Research team and Reflexivity

Prior to the study commencement, eleven participants were acquainted with ENI, none with KR. Participants were informed about the background and aims of the study before participating.

Both interviewers were researchers at the time of the study, residing in Germany. KR is of Austrian descent and identifies as female. She holds an MD, an MSc in Public Health and an MSc in Applied Epidemiology, and has training and experience in qualitative research methods including study design, semi-structured interviewing, and data analysis. She has completed a field epidemiology training programme and has received training in evaluation methods and evidence-informed decision-making in public health. She also has direct experience working within a ministry of health alongside national governmental decision-makers. ENI is of Bangladeshi descent, grew up in Germany, and identifies as male. He has a PhD in Physics of Biology and Complex Systems and an MSc and BSc in Physics, and has prior experience conducting Delphi-style expert consultations and infectious disease modelling for decision-making purposes.

Both researchers were educated primarily within European institutions – with ENI spending some time in Japan and KR in Gabon – and Western knowledge systems, with their respective disciplines oriented largely toward positivist epistemologies. While both have lived and worked outside high-income settings and believe this experience broadened their understanding of diverse health system and decision-making realities, they acknowledge that their high-income country and European cultural backgrounds inevitably bring a Westernised perspective to their understanding of health systems globally. They further acknowledge that their predominantly scientific training and work experience may shape their view of political decision-making, including a potential tendency to underestimate the inherent complexities of policymaking, the bounded rationality of decision-makers, and the limits of scientific evidence as a driver of policy. Throughout the study, both researchers remained conscious of these potential biases to reduce their influence on study design, data collection, and interpretation.

### Analysis

Recordings were transcribed in the language of the interview. All subsequent analysis, coding, and theme development was conducted in English. We employed thematic analysis (Braun & Clarke, 2006; Kiger & Varpio, 2020), with coding combining deductive and inductive elements. Deductive codes were guided by the research gaps, identified in our introduction – namely, the perspectives of knowledge producers, users and brokers on the successful use of modelling evidence in decision-making and the prevention of associated risks, the structural factors shaping modelling-to-policy pathways, differences between emergency and routine decision-making contexts, and national and international modelling-to-policy systems. We used inductive coding to capture experiences and events not anticipated by the deductive framework and to develop a comprehensive understanding of modelling-to-decision-making pathways. We conducted the analysis using MAXQDA 24.

Our analytical process moved iteratively from open coding of individual transcripts toward more focused coding, as recurring patterns across interviews were identified and consolidated. The resulting codes were then grouped into broader candidate themes, which the team reviewed, discussed, and refined collaboratively to ensure they coherently captured patterns across the dataset. Through this iterative process of comparison and refinement, themes on successful modelling evidence perceptions, risks, and factors influencing modelling evidence use emerged which clustered around the domains of policy relevance, model quality and communication and interaction. In a second analytical step, we applied a backward-mapping-informed approach to structure these findings into a multi-level theory of change. (Roberts & Khattri, 2012; Yearwood, 2018) Starting from the pathway outcomes, i.e. modelling evidence use, we worked backwards to group influencing factors across three logical levels accompanied by the proposed mechanisms linking them: the factors directly influencing these outcomes, then the conditions enabling those factors, and finally the system-level, structural inputs giving rise to those conditions. Tables 1 and 2 in the supplementary files present the identified factors and proposed mechanisms in full.

**Table 1:** Participant characteristics. “KP” is shorthand for knowledge producer, such as modeller, “KB” for knowledge broker, such as an advisor with technical background, and “KU” for knowledge user, such a decision-maker.

| Participant N° | Gender | Role | Location | Modelling-to-policy context |
| --- | --- | --- | --- | --- |
| 1 | Male | KP | HIC; Europe | International |
| 2 | Male | KP | HIC; Europe | Both |
| 3 | Female | KP | HIC; North America, Europe | Both |
| 4 | Male | KP; KB | HIC; North America, Europe | Both |
| 5 | Female | KP | Lower MIC; Africa | National |
| 6 | Female | KB; KU | HIC; Australia/Oceania | National |
| 7 | Male | KB; KU | HIC; Europe | International |
| 8 | Female | KP | HIC; Europe | International |
| 9 | Male | KP | HIC; Asia | National |
| 10 | Female | KP | HIC; North America, Europe | International |
| 11 | Male | KB; KU | HIC; Europe | National |
| 12 | Male | KP; KB | HIC; Europe | National |
| 13 | Male | KB; KU | HIC; Europe | National |
| 14 | Male | KP; KB | Upper MIC; Asia | National |
| 15 (with 16) | Male | KP; KB | Lower MIC; Africa | National |
| 16 (with 15) | Female | KP | Lower MIC; Africa | National |
| 17 | Female | KP; KB | HIC; Europe | National |
| 18 | Male | KP; KB | HIC; Europe | International |
| 19 | Female | KP; KB | HIC; Europe | National |
| 20 (with 21) | Female | KP | Upper MIC; Africa | National |
| 21 (with 20) | Female | KP | Upper MIC; Africa | National |
| 22 | Female | KP; KB | HIC; Europe | Both |
| 23 (with 24) | Male | KP | Upper MIC; Asia | National |
| 24 (with 23) | Male | KP; KB | Upper MIC; Asia | National |
| 25 | Male | KP | HIC; North America | Both |
| 26 | Male | KP; KB | HIC; Europe, Asia | Both |
| 27 | Female | KB | Lower MIC; Asia | National |
| 28 | Male | KB; KU | LIC; Africa | National |
| 29 | Female | KP | Lower MIC; Africa | National |
| 30 | Female | KP; KB | Lower MIC; Asia | National |
| 31 | Female | KP | Lower MIC; Africa | National |
| 32 | Female | KP; KB | HIC; Australia/Oceania | National |
| 33 | Male | KB; KU | HIC; North America | International |
| 34 (with 35) | Male | KP | Lower MIC; Asia | National |
| 35 (with 34) | Female | KP | Lower MIC; Asia | National |

Due to capacity constraints, the majority of data coding was carried out by KR, contrary to the plan specified in the study protocol. ENI independently coded two transcripts, yielding a total of 76 codes. Codes were subsequently compared between coders to assess consensus and ensure completeness. Inter-coder agreement was high for code completeness: Only five codes generated by ENI were not captured in KR’s coding, suggesting that KR’s coding was largely comprehensive.

Participants did not provide feedback on the findings, but a preprint version of this manuscript was shared with those having stated interest.

## RESULTS

Between May and September 2025, we interviewed 35 key informants, including 16 knowledge producers (KP; modellers), 13 knowledge brokers (KB) of which 12 also had experience as knowledge producers, and 6 persons having both been involved in knowledge brokering and use (KU; executive decision-makers). The latter included heads of national public health institutes and persons employed in executive roles at national and international organizations in the field of public health. Twenty participants were based in high-income (HIC) and 15 in low-and-middle-income countries (LMIC), according to the 2025/2026 World Bank country classifications by income level (Metreau et al., 2025), on five continents (Africa, Asia, Australia/Oceania, Europe, North America), working in national (n=23), international (n=6), or both contexts (n=6). (Table 1)

The results are structured to follow our research questions and analytical workflow. We begin with participants’ perspectives on what constitutes successful modelling evidence use and potential associated risks, before turning to the factors that influence these outcomes, the conditions that enable them, and the system-level inputs that give rise to these pathways. We then report how these pathways differ between routine and emergency decision-making settings, and between national and international modelling-to-policy systems.

### Perspectives on Successful Use of Modelling in Decision-Making and Potential Risks

Participants held divergent views on what constitutes successful modelling-to-decision-making pathways and outcomes. Some participants, predominantly modellers, conceptualized success as the instrumental use of modelling evidence – that is, policy decisions directly based on and aligned with modelling results.

> **Quote 1:** *“So, the big success is that now the country has revised its control strategy based on the model outputs. And that has been adopted at the national level, and WHO has also looked into the work that has been done and referencing to what the modelling has been able to achieve […]. That’s a big success.”* (Participant 5 – KP, national context)

However, other participants – mostly knowledge brokers and users, but also some modellers – challenged this framing, recognizing the inherent complexity of decision-making processes, particularly in political contexts where multiple competing factors exert influence. These participants perceived success as the meaningful consideration and comprehension of modelling evidence by decision-makers, even when final policy choices diverge from recommendations implied by model results. This perspective acknowledges that while scientific advice aims to inform decision-making, democratic policymaking must consider a variety of factors other than scientific evidence.

> **Quote 2**: “*So, success for me is if the politicians communicate about the evidence accurately. Sometimes evidence points in favour of saying: “We need to go into that direction.” And then politics - So, then ideology is used to interpret certain evidence and saying, like: “I am looking at the evidence but I believe that the path forward is something else.” That is – for me – that is how politics works. That is also a good thing. Because otherwise politicians just implement what scientists are saying. And then it is not policy anymore. So, success for me is when a politician accurately interprets the evidence at hand and then decides based on basically what democracy wanted.*” (Participant 12 – KP and KB, national context)

Despite these differing conceptualizations of successful modelling evidence use, participants converged on a critical concern: the risk of decisions based on misunderstood, overly trusted, not appropriately contextualized modelling evidence, leading to policies not justified by the evidence or misaligned with the actual epidemic situation or public health goals such as an inappropriate use of resources. Participants reported instances where decisions based on misunderstood modelling evidence further led to losing trust between modellers and decision-makers and thus sustainability. We identified two primary risk factors in modelling-informed decision-making. First, **inadequately specified models** – those built on inappropriate assumptions or lacking proper contextualization for the decision at hand – could generate misleading results. Second, **failures in communication** between modellers and decision-makers, alongside a limited understanding of model uncertainty, could result in outputs being misinterpreted or overly trusted. Quote 3, from a modeller’s perspective, illustrates both risk factors: an overly trusted, prestigious evidence source combined with evidence users lacking the technical understanding to judge model quality, given the perceived complexity of this type of evidence.

> **Quote 3:** *I think there’s a number of fairly famous examples of Ebola models that went wrong and models that were … mis-specified. They had errors in the way the models were made. And I think … one of those models had a lot of impact in terms of drawing attention because it was projecting a lot of cases. I feel like that sort of thing works only once or twice when the policymakers are new to models, … and that was the case maybe in 2015-ish, or 2014, in Ebola. Many hadn’t seen models. And then I think it was very hard at that time for a policymaker that also didn’t know what was going to happen, but had maybe their idea to say: “Oh, your model that comes from well-known people with a sort of mathematical structure and seems very objective from outside, but I can’t because I don’t have the skills to look at it from, to me, it’s a black box.” Then, then it’s very hard to say: “Oh, this is wrong. It’s not what’s going to happen.” (Participant 18 – KP and KB, international context)*

Knowledge users echoed this concern, highlighting communication failures arising when modellers did not adequately or transparently convey the uncertainties and limitations of their outputs. One knowledge user, reflecting on how uncertainty was communicated to them, described considerable variation in practice:

> **Quote 4**: *Certainly, some are quite correct, both in publication and in naming the limitations, doing sensitivity analyses, where uncertainty factors might be, and so on and so forth. And some are a bit more casual about it, I’d say, and like to sell it as truth, … I think I’ve actually seen both. … I think overall most of them do say what their models can and cannot do. Whether that’s always understood when they express it in such mathematical terms, that would be a somewhat different question.* (Participant 11 – KB and KU, national context)

This variation in transparency compounds a further risk: that communicated projections, or central estimates are taken at face value, particularly by non-technical audiences, without due regard for the uncertainty inherent in modelling – a risk that is especially pronounced when little is yet known about the disease being modelled. Reflecting this concern, another decision-maker described a marked preference for relying on solid empirical evidence over modelling outputs wherever possible:

> **Quote 5**: *I would rather have facts, I would rather have solid evidence than modelling outputs. … I would rather see the actual figures I wouldn’t use modelling alone because it’s not the truth, it’s not the evidence. It’s based on what might occur, or what could occur. … I would not base any policies, other than precautionary measures in some instances, on that. … So, you know, modelling has its place, it’s a very important public health tool, it can project what might be the maximum, what might be the minimum, if you have the right input … But if it’s something that there is not much evidence about and modellers are just sort of suggesting what might happen, I don’t think that’s good. And I think once a model is done, policymakers … need to understand the limitations of modelling and these are only estimates, and often times it doesn’t occur, it’s just taking this fact, here is what will happen.* (Participant 7 – KB and KU, international context)

In summary, participants perceived success in modelling-for-decision-making as either the understanding and consideration of modelling evidence, or as its instrumental use in decision-making. Yet, they converged on the need to prevent risks to modelling-informed decision-making.

### A Three-level Framework to evaluate Modelling-for-Policy Pathways and Systems

To map the pathways and mechanisms underlying perceived successful modelling evidence use, and to link these to system-level factors, we used a backward mapping-informed approach based on participants’ accounts: starting from the factors that directly influence perceived successful use and mitigate associated risks (Level 1), we then identified their enabling conditions (Level 2) and, in turn, the systemic enablers (Level 3) that give rise to these conditions. We propose causal mechanisms linking each level, describing a theory of change of how system-level inputs can increase the utility of modelling for policy while mitigating its risks within a framework – that can be tested in future work. Figure 1 illustrates the structure of the framework. The following sections describe each level in turn.

**Figure 1:**
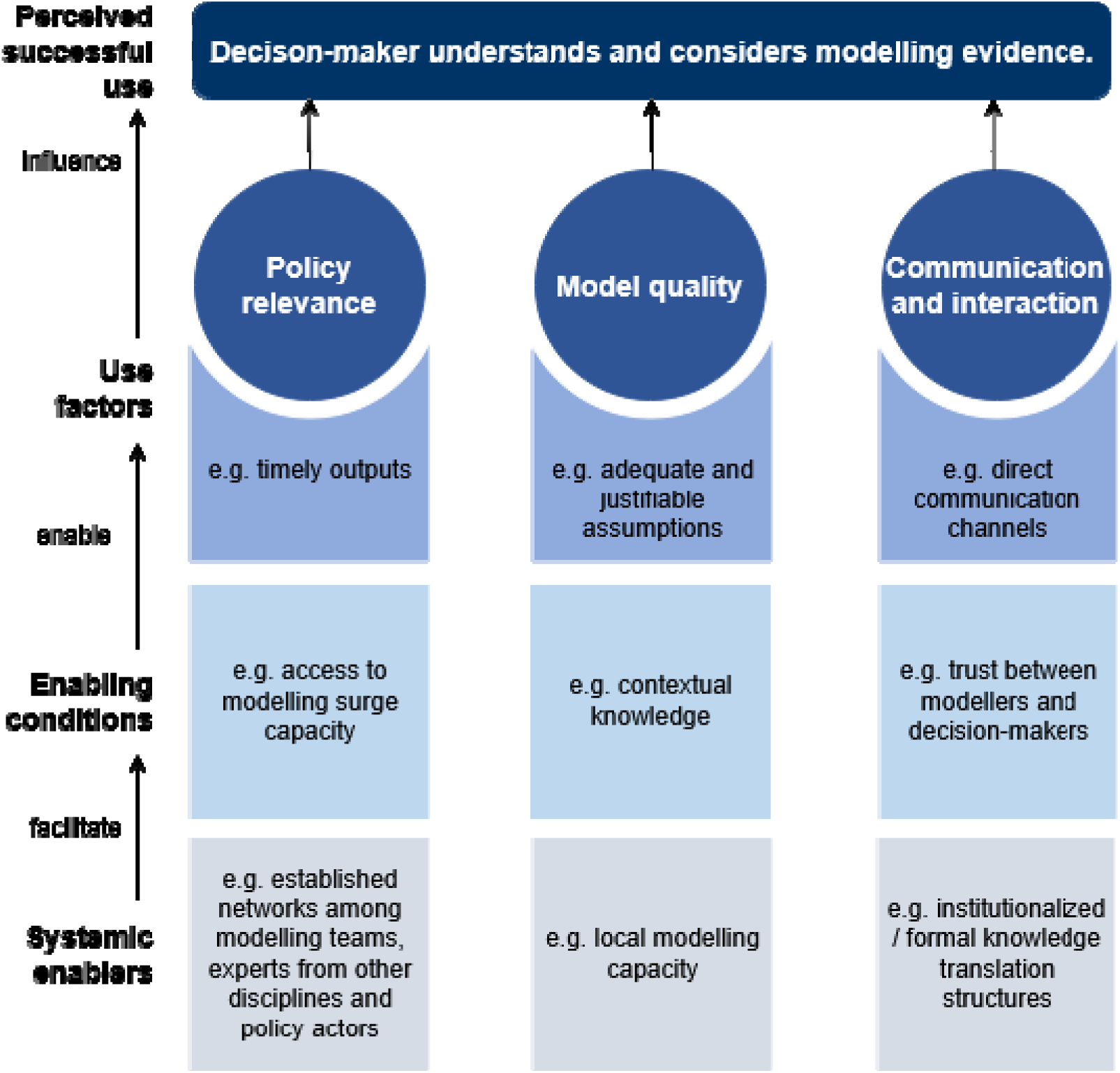
Structure of the three-level evaluation framework on modelling-for-policy pathways.

### Level 1: Factors influencing the Perceived Successful Use of Modelling Evidence

Despite participants’ divergent understandings of what constitutes meaningful modelling evidence use – whether as the accurate understanding and consideration of evidence, or as its instrumental application in decision-making – we were able to identify factors that influence use under both understandings. Since the accurate understanding and consideration of evidence is a necessary precondition for its instrumental use, any factor influencing how decision-makers engage with modelling evidence will also influence its instrumental use. Our analysis revealed that these factors clustered around three interdependent domains: policy relevance, model quality, and communication and interaction.

**Policy relevance** encompasses the alignment between modelling outputs and the actual needs of decision-makers. Models developed in isolation from policy contexts – however technically sophisticated – were seen as unlikely to usefully inform decisions. Not every policy question, moreover, can be meaningfully addressed through modelling; for outputs to be policy relevant, the underlying question must be amenable to modelling, and results must be available within the decision-making timeframe. Models that were co-created with decision-makers or knowledge brokers, and that could be adapted and reused as policy needs evolved, were perceived as more relevant, as they more accurately captured decision-makers’ needs.

**Model quality** refers to the technical and methodological soundness of the modelling approach. Participants noted that model quality alone does not guarantee utility for policymakers, but poor quality was seen to undermine credibility that can lead to losing trust and disrupt future engagement while risking generating misleading results. Factors within this domain include adequate and justifiable model assumptions for the given purpose and context, peer-review processes, and a diversity of modelling approaches addressing a given policy question, which allows structural uncertainty to be challenged. Two further factors sit within model quality while relating closely to policy relevance: holistic modelling and equity considerations. Holistic modelling integrates a diversity of perspectives – from other disciplinary experts such as behavioural scientists or economists, as well as those affected by the policy decision – capturing real-world complexity in ways that can enhance both model quality and policy relevance. Equity considerations within the modelling process reduce the risk of policy being informed by biased results that overlook the differential impacts of decisions across population groups.

**Communication and interaction** emerged as critical bridging factors between relevant, high-quality models and their use in decision-making. This domain spans both the clarity and accessibility with which results – and particularly uncertainty – are communicated and tailored to decision-makers, which emerged as key to preventing misunderstandings, and the relational dimension of ongoing dialogue between modellers and decision-makers. Factors here include direct communication channels between modellers and decision-makers that minimise loss of nuance in translation, iterative and bi-directional interaction enabling feedback and refinement, and sustained engagement that builds a foundation for long-term relationships and future modelling-for-policy collaboration.

Figure 2 lists all the identified factors. Table 2 in the supplementary files provides detailed descriptions of the factors within each domain that we identified in our analysis, along with their intersections. We do not claim that all factors must be present for modelling evidence meaningfully informing decision-making; the relative importance of individual factors remains to be established as this framework is applied to specific decision-making settings.

**Figure 2:**
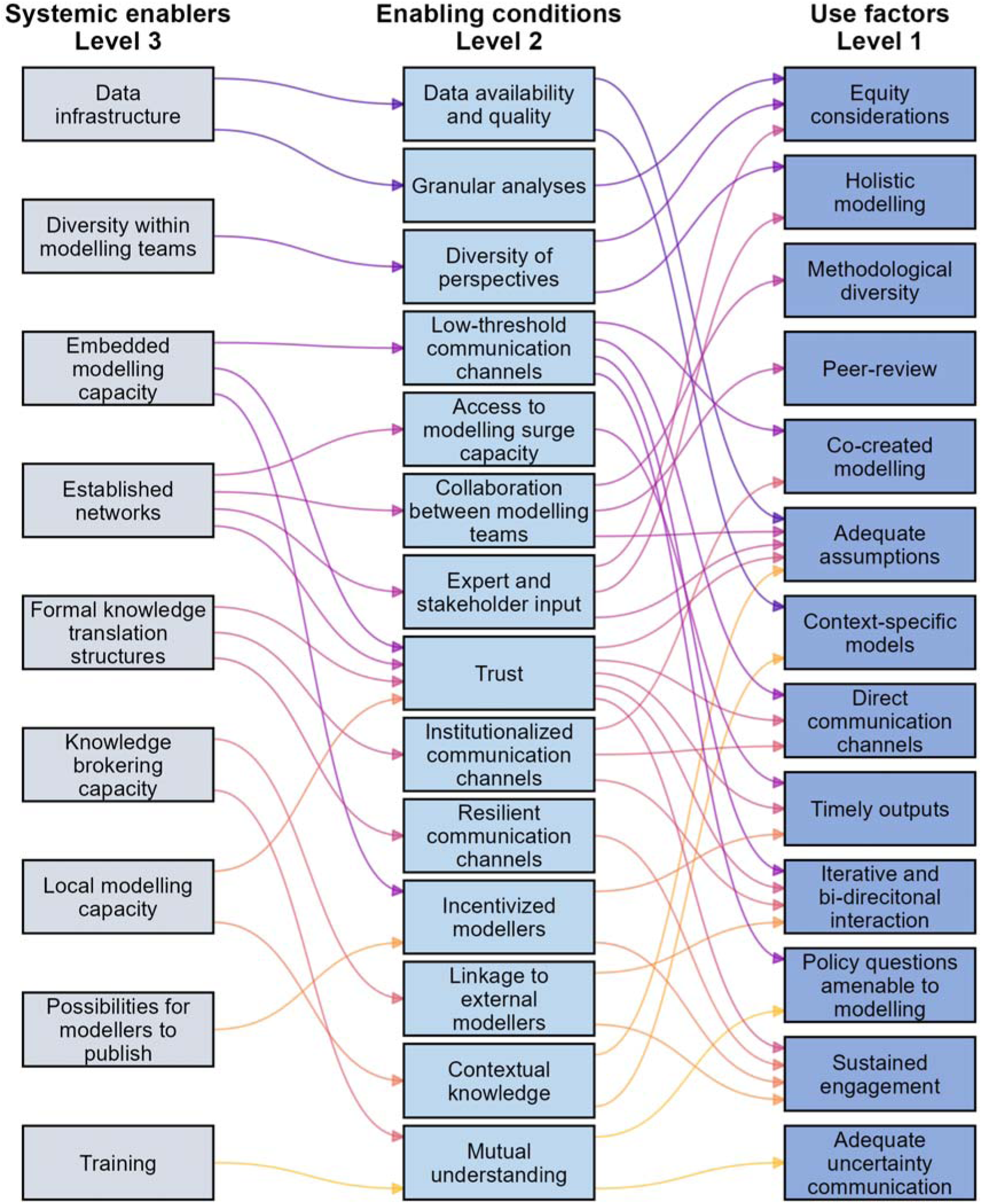
Identified factors and pathways within the three-level framework. Figure 2 provides and overview of all identified factors (boxes) within each level of the framework and their linkages (arrows).

### Level 2: Enabling Conditions

Based on participants’ accounts of preconditions and applying a backward-mapping-informed approach, we were able to identify a second level of factors that sit between the factors directly influencing modelling evidence use (Level 1) and the structural, system-level inputs described below (Level 3). We term these enabling conditions: they represent intermediate states or outcomes within a modelling-to-policy system that our data suggested were relevant for Level 1 factors to be present, but that were not themselves directly actionable structural inputs. Several enabling conditions were found to link to more than one Level 1 factor, and in turn to more than one Level 3 systemic enabler (Figure 2); a detailed account of these proposed relationships is provided in Table 3 in the supplementary files.

The conditions we identified are as follows.

Modellers’ **contextual knowledge** enables the creation of models that are specific and relevant to the decision-making setting they aim to inform and supports adequate assumptions for that context – thereby enabling factors related to both model quality and policy relevance.

The **quality and availability of data** relevant to the decision-making context, and the **granularity of analyses** performed, are conditions relevant to model quality and policy relevance alike: they support adequate assumptions and context-specific models, and the consideration of equity questions within the modelling process.

**Collaboration with other modelling teams**, together with **expert and stakeholder input** and a **diversity of perspectives** within the modelling team, enable factors within the model quality domain. Diverse perspectives support holistic modelling and the consideration of equity questions, while collaboration between modelling teams facilitates peer review and can increase methodological diversity where teams employing different approaches are involved.

**Trust** and **mutual understanding** between modellers and decision-makers enable more direct and iterative communication, allowing for feedback and bi-directional exchange. Trust can further motivate data sharing with modellers – particularly relevant for those not employed by government. Thereby it supports timely outputs and can motivate sustained engagement for future collaboration. Mutual understanding of policy needs, and of what modelling can offer, increases the likelihood that the questions asked are amenable to modelling and that uncertainty is communicated and understood adequately.

**Low-threshold and resilient communication channels** further enable factors related to interaction, such as more direct and iterative communication.

**Access to surge modelling capacity** enables the production of timely outputs where policy needs and time constraints shift rapidly, such as in emergency situations.

**Incentives for modellers** to produce policy-relevant outputs, rather than prioritising, e.g., academic outputs, further support the production of timely outputs that meet policymakers’ needs, as well as sustained engagement between modellers and decision-makers.

### Level 3: Systemic Enablers

At the third level, based on participants’ accounts, we identified structural, system-level inputs that linked to the enabling conditions described in Level 2 (Figure 2). We term these systemic enablers: unlike enabling conditions, they represent tangible features of a modelling-to-policy system. As with Level 2, several systemic enablers were found to link to more than one enabling condition and, through them, to more than one Level 1 factor. Table 3 in the supplementary files provides a full account of these proposed pathways. The systemic enablers presented here reflect what emerged from our data; additional systemic inputs may exist that were not captured in our dataset.

We identified the following systemic enablers:

**Local modelling capacity** facilitates context-specific models and adequate assumptions through the contextual knowledge that local modelling teams hold regarding disease dynamics, health systems, and governance environments. It can also foster trust between modellers and decision-makers, although depending on local modellers demonstrating sufficient technical expertise, since internationally renowned institutions may otherwise be perceived as inherently more credible by decision-makers.

**Embedded modelling capacity** (modelling teams situated within or closely tied to decision-making institutions) facilitates and low-threshold communication channels and trust between modellers and decision-makers. This supports direct, iterative, bi-directional interaction, co-created modelling, and sustained engagement. In smaller countries, the limited number of stakeholders involved can produce similar relational proximity even without in-house capacity. Embedded modellers may also face stronger incentives to prioritise policy-relevant, timely outputs given their institutional funding and accountability.

**A well-functioning data infrastructure** that encompasses timely sharing of comprehensive, reliable data facilitates adequate assumptions and context-specific models by enabling more accurate parameterisation. Where data infrastructure provides comprehensive data – including information on specific population subgroups – it enables granular analyses for equity considerations to be incorporated into modelling outputs.

**Knowledge brokering capacity** facilitates mutual understanding between modellers and decision-makers, which in turn supports adequate communication of uncertainty and the formulation of policy questions that are amenable to modelling. Brokers with proximity to decision-makers can also link them to external modellers, facilitating iterative interaction and sustained engagement that external teams might otherwise struggle to establish independently.

**Formal knowledge translation structures** – such as advisory boards or policy dialogue – facilitate trust between modellers and decision-makers through repeated, structured engagement, enabling iterative interaction, co-creation of models, and more direct, informal communication channels. By possibly increasing the resilience of these relationships to changes in government, formal structures can also support sustained engagement over time – particularly for external or academic modellers not embedded within decision-making institutions.

**Training** for modellers **on communication and policy needs** and **decisionmakers’ familiarization with modelling techniques** facilitates mutual understanding by improving modellers’ ability to communicate uncertainty adequately, and by enabling decision-makers to assess what questions a model can and cannot answer, interpret outputs critically, and appreciate caveats and uncertainty – both reducing the risk of misunderstandings.

**Established networks** among modelling teams, other disciplinary experts, stakeholders and policy actors facilitate several conditions. Collaboration between modelling teams supports peer review, adequate assumptions, and methodological diversity, allowing structural uncertainty to be appraised when multiple teams address the same question. Expert and stakeholder input broadens model development to produce more holistic, equity-sensitive outputs and improves the quality of assumptions. Established networks can also provide access to modelling surge capacity, supporting timely outputs in fast-moving or emergency contexts, and foster trust through continuous interaction.

**Diversity within modelling teams** (across gender, ethnicity, and geographic or cultural background) facilitates diversity of perspectives in model development, increasing the likelihood of holistic and equitable modelling outputs.

**Possibilities for modellers to publish** facilitate incentivised engagement, particularly for academic modelling teams, thus motivating timely outputs and sustained engagement with decision-makers – for instance through agreements that protect publication rights even where decision-makers retain ownership of underlying data.

### Funding as a Cross-cutting Systemic Enabler

We found funding to be a cross-cutting input, i.e. being a pre-requisite for several systemic enablers such as a well-functioning data infrastructure, modelling and knowledge brokering capacities. Continuous funding supports sustained engagement between modellers and decision-makers and enables modellers to focus on policy-relevant work. While government-employed modellers mostly have this financial sustainability, academic modellers often depend on third-party funding which – if prescriptive – can limit flexibility in choosing relevant health policy topics. Non-prescriptive funding can enable academic modellers to focus on policy relevant work. Discontinuation of funding can lead to a loss of research capacity and engagement. Quote 6 illustrates this experience of an academic modeller after the COVID-19 pandemic, where reduced government interest led to funding cuts, forcing a shift in research focus.

> **Quote 6**: „*Personally, I haven’t received any more funding And that means my research in this area is over. These are long-term decisions. If I reduce my expertise in this area now, it means I no longer have the group for it. Period. Then I won’t start a new topic unless there is really long-term funding for it. Then I’ll move on to another topic. … 90 percent of my budget comes from third-party funding. So, in that respect, research funding largely determines what we end up researching. And if I no longer have the people in the group, I’m not going to rebuild it from scratch if I don’t know that it will be long-term.*” (Participant 19 – KP and KB, national context)

Participants further highlighted that funding should be local and independent of international aid, as donor funding can be volatile and bear the risk to leave decision-makers without access to modelling capacity. This is illustrated in Quote 7 arguing for more local modelling capacity to ensure sustainability.

> **Quote 7**: “*First of all, my vision was the Southern institutions working locally should also at some point make sure that they are sustaining themselves independently of external funding. Like were they hired by or is it embedded in the Ministry of Health or is it a service that is provided by local institution paid by the Ministry of Health? – However, the constellation could be locally. But that it sustainable. We seek funding from international aid. But like especially in the last few months and few weeks even we see: There will be an end to this. … So, we need to converge to something that is more sustainable. And for me that starts with moving this capacity in the South.”* (Participant 8 – KP, international context)

### Preventing Risks to Modelling Evidence Use: Uncertainty Communication

We found the communication and understanding of uncertainties around modelling outputs to be critical in preventing risks for decision-making informed by modelling arising from communication failures that can occur even if model quality is high. Modellers and knowledge brokers highlighted challenges in getting nuances across and the risk of only the central estimate getting picked up by decision-makers. Our analysis identified several factors that facilitate uncertainty communication, understood here as modellers communicating transparently and decision-makers understanding correctly. These include tailoring messages to the audience, maintaining iterative and bi-directional interaction that allows for follow-up questions and refinement, and using direct communication channels that minimise the risk of nuance being lost in translation.

Iterative, direct, and bi-directional engagement between modellers and decision-makers proved especially important – allowing modellers to explain assumptions and evolving outputs and giving decision-makers space to ask clarifying questions. Our analysis suggests that such interaction can be actively enabled through systemic factors such as institutionalized knowledge translation structures, e.g. formal advisory boards, modellers embedded within decision-making institutions, and the inclusion of local modellers when working with international teams. These inputs tend to facilitate enabling conditions for iterative, direct and bi-directional interaction such as relational proximity and trust between modellers and decision-makers, as well as low-threshold and institutionalized communication channels. In direct communication formats, group-based discussions involving multiple modellers were similarly perceived as more effective than one-to-one interactions, as they reduce the risk of selective uptake and encourage greater transparency about assumptions and limitations. Formal advisory structures were described as fostering exactly this kind of trust-based, transparent exchange. One modeller contrasted the dynamics of small-group advisory settings with one-to-one communication:

> **Quote 8**: *“I think this small-group aspect, where colleagues are sitting right there listening in, is much, much better than one-to-one communication, because there can be a tendency for politics to simply seek out experts who are easy to understand … which often means [that experts] who don’t communicate the uncertainty are more often the ones approached. … We had that relationship of trust, so that even absurd or otherwise unsayable things could, so to speak, be said.“* (Interview 17 – KP and KB, national context)

A decision-maker described how such structures had become embedded over time through dedicated institutional funding, creating a standing point of contact between modellers and policymakers:

> **Quote 9:** *“There are, of course, all kinds of expert panels that meet, and so on. There are always modellers in there too, so you get into conversation with them. … and, I’d say, the whole thing became institutionalized over time through [governmental] funding that supported this modelling network … and that’s actually a really good opportunity … And that’s another network where you always have someone you can turn to.“* (Participant 11 – KB and KU, national context)

Institutionalized knowledge translation structures do, however, risk creating blind spots. By channelling access to decision-makers through established structures, they may exclude methodological approaches and modelling voices that sit outside them – narrowing the evidence base and potentially limiting how comprehensively policy questions are addressed.

Embedding modellers within or close to decision-making institutions was similarly described as enabling the kind of sustained, bi-directional interaction allowing for feedback. One modeller supporting outbreak responses from a geographically distant location contrasted the effect of having an embedded local point of contact with more distant arrangements:

> **Quote 10:** *“There is no real clear feedback mechanism for knowing what was useful and what wasn’t. … I think what helps is if we have an in-country partner, or somebody who is seconded to the operating centre. That’s a very useful connection. …, because if you are not really embedded with the other team, sometimes it’s just separate teams passing information over without really knowing where it goes.“* (Participant 1 – KP, international context)

Another modeller linked this proximity directly to the consistency of feedback received:

> **Quote 11:** *“I think we received feedback as often as we sought feedback. It’s about keeping this two-way communication open. … the institutional setting where policymakers were simply close [mattered] – not everyone has the luxury of these intense, constant interactions with the policymakers they are supporting. Within [name of institution], we have that luxury – many of the decision-makers are right next door, or long-term colleagues.“* (Participant 3 – KP, national and international context)

The format in which uncertainty is communicated matters considerably and depends on the audience. Scenario-based outputs were widely seen as more accessible and useful for non-technical audiences than point estimates or probabilistic expressions. In contrast, central estimates and confidence or prediction intervals were perceived to be more difficult to get across correctly and bear a higher risk of loss in nuance and selective interpretation – especially when audiences focus on single values of interval bounds. Such simplification risks eroding trust if central estimates later prove inaccurate. This is illustrated in Quote 12 by a participant acting as knowledge user with technical literacy and knowledge broker to political levels.

> **Quote 12**: “*And of course, the weakness is that many of these estimates are uncertain and for each of the estimates there is a broad confidence limit and when you have then a lot of different estimates factored into the same mathematical model and all the uncertainties are multiplied then of course at the end of the day, the confidence of your mathematical models are quite wide. I think as a scientist you can deal with that and you can say: “Okay, but probably I mean the central estimate is what we see as the most likely, but it could be anything from this to this.”… But for politicians and for the public, that is much harder to digest. But the problem is if you then only communicate the central estimate that’s often not a good position to be in because if then it turns out the central estimate wasn’t a complete correct estimate, then people would say you were wrong again*.” (Participant 13 – KB; KU, national context)

Decision-makers’ technical knowledge and the presence of skilled knowledge brokers – capable of bridging technical and policy worlds – were identified as important facilitators of accurate uncertainty communication and interpretation. Their technical literacy was perceived as facilitating the interpretation of distributions, ranges, and assumptions. For time-pressured or non-technical audiences, simpler visual representations and ranges were perceived more useful – by modellers mostly - than probabilities. However, such simplification carries the risk of losing critical nuance. Skilled knowledge brokers understanding both technical detail and policy needs, are well positioned to mitigate such risks by translating uncertainty in ways that are meaningful and actionable to decision-makers. Additionally, standardized visual and linguistic conventions – such as those developed by the Intergovernmental Panel on Climate Change – were highlighted as promising models for navigating this complexity as outlined in Quote 13.

> **Quote 13**: “*So rather than presenting that central estimate, presenting the distribution, and I think there’s much better visualizations for doing this now that are visually appealing, informative, but still capture that uncertainty. I also think there’s a lot of improvement in the language around talking about uncertainty, partly driven by some of the work in climate modelling. You know, the IPCC (note: intergovernmental panel on climate change) reports have got nice ways of translating uncertainty into the language that they use. And I think that kind of standardization is nice*.” (Participant 22 – KP; KB, national and international context)

### Modelling-to-Policy Pathways in Different Settings

### Time-Constrained Settings vs. Routine Decision-Making

When comparing emergency settings – such as outbreak responses – with more routine decision-making contexts, such as national immunization planning, our analysis revealed that the relative importance of certain factors varied: Timeliness emerged as a distinctly more pressing concern during emergencies. In such settings, participants faced an explicit trade-off between model quality and the timeliness of results, where outputs that arrive too late to inform decisions carry little policy relevance regardless of their technical rigour. This tension is captured by one knowledge producer operating across national and international contexts in Quote 14.

> **Quote 14**: “*Yeah, for sure, depending on the context, sometimes 80% good is enough and going for the remaining 20% is really a bad decision because it impacts on the timeliness of your outputs and whether they can actually influence decision-making or not. So definitely … during outbreak responses, this is something that you are always negotiating with*.” (Participant 2 – KP, national and international context)

Across our data, the risks to modelling-informed decision-making emerged as particularly pronounced in time-constrained settings, such as emergency contexts. The urgency of decision-making in these settings often precludes triangulation with other evidence types – such as randomized controlled trials or alternative study designs – that cannot be produced at the speed decisions demand. As a result, modelling evidence may carry greater singular weight in shaping decisions, as one modeller described his perceptions:

> **Quote 15**: *” … sometimes they would have to make a decision based only on this analysis. They were like: okay, we understand that you can’t project something six months ahead of time, but this is all we have to work with, so we just have to go with these numbers. So, they would have an understanding that this is a highly uncertain result, but at the same time they would also say there is nothing else we have to work with, so we are just going to use this central estimate.“* (Participant 1 – KP, international context)

Our analysis suggests that pre-established networks between modelling communities and decision-makers can help minimize the trade-off between model quality and timeliness by enabling rapid evidence production in time-constrained settings, sustaining relationships at both national and international levels and allowing additional capacity to be mobilized quickly when needed. In the absence of such networks, participants described the matching of urgent policy questions to available modelling capacity as happening largely by chance, dependent on personal connections rather than a systematic process – some noted the risk of institutions being “left behind” simply because they lacked an established relationship to call on. Beyond mobilizing capacity faster, participants suggested that networked, multi-group arrangements could also enhance model quality by enabling forms of rapid peer-review that the urgency of emergency settings would otherwise foreclose. One modeller contrasted the well-established, peer-reviewed validation pathways available for routine policy questions with the gap facing urgent, time-critical ones, and proposed networks like rosters of modelling groups as a way to bring comparable scrutiny to fast-turnaround settings:

> **Quote 16**: *“For things that are more long-term policy questions… what we have now as a system, with peer-review – I guess you probably know about SAGE, there is a sort of consortium that grades model outputs for SAGE. If you want SAGE to use your model, you need to get positive advice from that team… that, for me, is a very good process, and it’s already quite well settled. But where right now it’s not satisfactory is the urgent questions…, where you need to decide in a very short time and there is no journal peer-review possible. So there I’d see some sort of platform where different modelling groups submit their answers as a good future [model].“* (Participant 18 – KP and KB, international setting)

### International Modelling-to-Decision-Making Systems

Across our data, we observed considerable diversity in how modelling-to-decision-making systems are structured – particularly in terms of where decision-making bodies and modelling teams are located and how they interact. Decision-making itself may occur at local, national, or international levels, the latter encompassing international governance bodies such as the WHO as well as NGOs operating globally within processes such as outbreak responses. Importantly, modelling capacities are not always co-located with the decision-makers they inform: we identified settings in which national or international decision-making is supported by modelling teams based elsewhere, sometimes alongside local modelling capacity and sometimes without it.

Established international networks of modellers, intermediaries, and decision-makers were found to facilitate rapid engagement and the scaling of capacity – assets that are particularly valuable in outbreak situations or in settings with limited local resources. However, relying on geographically removed modelling teams carries heightened risks to modelling-informed decision-making – both through a lack of contextualized model specification and communication challenges inherent in geographically and institutionally distant relationships with increased risk of losing nuance.

Participants highlighted that incorporating local modelling capacity can mitigate both risks. Local modellers bring contextual knowledge that supports more appropriately specified models, and their proximity to decision-makers can enable more direct, iterative, and trust-based communication. Relying exclusively on international modelling capacity therefore not only raises concerns about the long-term sustainability of modelling systems – as outlined above – but also introduces risks to modelling-informed decision-making.

## DISCUSSION

This study explores perceptions of modellers, knowledge brokers and decision-makers on the use of infectious disease modelling evidence in public health decision-making and its influential factors. It developed a three-level framework mapping pathways and mechanisms how structural, system-level factors facilitate modelling evidence use in public health decision-making (Figure 1). Participants diverged in how they conceptualised meaningful use – as accurate understanding and consideration of evidence, or as its instrumental application in policy – yet converged on a shared concern: that misunderstood, overly trusted, or poorly contextualised modelling evidence can present risks to modelling-informed decision-making and erode trust between modellers and decision-makers. Drawing on this, we identified factors directly influencing modelling evidence use across three domains (policy relevance, model quality, and communication and interaction), traced these back to enabling conditions, and further to systemic facilitators – including local and embedded modelling capacity, data infrastructure, knowledge brokering capacity, formal knowledge translation structures, established networks and funding – that could be targeted through interventions. Adequate and transparent uncertainty communication emerged as a particularly important to prevent risks to modelling-informed decision-making arising from communication failures. The relative importance of the identified factors varied across settings: emergency contexts heightened trade-offs between timeliness and model quality, while reliance on geographically distant modelling teams introduced additional risks that local capacity and sustained networks helped to mitigate.

### Diverging Perceptions of Successful Modelling Evidence Use

A central finding of this study is the divergence in how participants conceptualized successful modelling evidence use in decision-making. Participants who were predominantly modellers tended to define successful use as policy decisions directly informed by and aligned with model outputs – while others, including those with experience spanning brokering and decision-making roles, adopted a broader understanding: that the consideration and accurate interpretation of modelling evidence by decision-makers constitutes successful evidence use, even when final decisions diverge from model recommendations.

Within the evidence-use literature, the conception of evidence directly translating into policy decisions corresponds to what is termed ‘instrumental use’. (Weiss, 1979; Weiss, 1980) Beyond this, various more indirect forms of evidence use have been described – such as conceptual use, where evidence shapes thinking or reframes problems without directly determining decisions – that may be less tangible to knowledge producers but are nonetheless valuable. (Oliver et al., 2014; Weiss, 1980) Empirical work suggests that these indirect forms of use are in fact far more prevalent than instrumental use, particularly when issues are complex, consequences are uncertain, and decision-making involves multiple actors. (Weiss, 1980)

This finding resonates with the argument that the ‘linear model’ of evidence-to-policy misrepresents how policymaking operates. (Cairney, 2016; Oliver, 2022) Democratic policymaking involves the negotiation of values, competing interests, institutional constraints, and political considerations, and evidence is one input among many. (Cairney, 2016) Our data reflect this reality: Decision-makers, knowledge brokers, and also some modellers we interviewed recognized this complexity as inherent to the policy process rather than as a failure of it.

The tension between these diverging success perceptions is well documented in the broader evidence-use literature, which has long observed that researchers’ expectations of instrumental use may conflict with the complex realities of how decision-makers engage with evidence. (Greer, 2022; Oliver, 2022) Modellers who default to instrumental use as their benchmark risk underestimating their own influence: where modelling evidence is shaping the cognitive landscape of decision-makers without directly determining decisions, this impact may go entirely unrecognized. This matters not only for how modellers assess their own contribution, but also for the sustainability of modelling-to-policy engagement more broadly. If modellers consistently perceive their work as unsuccessful – because it does not visibly and directly determine decisions –, this may generate frustration and disillusionment that undermines continued investment in the relationships between modellers and decision-makers.

Further, when modellers are incentivized – consciously or not – to maximize the easily visible policy impact of their work, i.e. strive for instrumental use, they may be tempted to present results with greater certainty than is warranted, or to frame outputs in ways that are suggestive rather than genuinely clarifying. This risk is particularly pronounced for modelling evidence, given that models can be parametrized in ways that steer results toward expected or desired outcomes. (Pepin et al., 2025) Saltelli et al. warn explicitly that modellers must not project more certainty than their models deserve, and that predictions risk serving certain political agendas rather than genuine deliberation. (Saltelli et al., 2020) If decision-makers perceive that modellers are pursuing an agenda or overselling their results, the consequent erosion of trust may damage not only individual relationships but the broader legitimacy of modelling evidence in policy. However, the drive for policy relevance might also have positive consequences: modellers oriented toward instrumental impact may invest more in understanding the decision context, engaging stakeholders iteratively, and producing outputs that are responsive to policy needs – factors that our own data identify as important success enablers.

### Preventing Risks of Modelling-for-Decision-making

Despite diverging perceptions of successful modelling evidence use, participants in this study converged on a critical concern: the existence of potential risks to modelling-informed decision-making. These risks are conceptually distinct and can arise independently of one another. Decisions may be informed by modelling evidence that is itself of poor quality – for instance, built on inadequate or poorly contextualised assumptions – regardless of how well that evidence is subsequently understood or communicated. Separately, decisions may be informed by modelling evidence that is sound but misinterpreted by those using it, for example through a failure to appreciate the uncertainty surrounding model outputs. And decisions may be informed by modelling evidence that is misleadingly presented, whether intentionally or not, irrespective of the underlying quality of the model itself. We do not seek to judge which of these risks is more consequential, nor whether the resulting decisions are themselves better or worse, rather, our concern is with the conditions that give rise to each of these distinct risks, as all three can carry consequences not only for the immediate decision but for the long-term sustainability of modelling-to-policy relationships through eroding trust.

Two primary risk factors emerged from our data: inadequately specified models and communication failures. Both pathways are reflected in the broader literature on modelling to inform decision-making, which has described model misspecification (James et al., 2021; Moran et al., 2025; Rhodes et al., 2020; Van Damme et al., 2020) or the inadequate communication of results as risks to modelling-informed decision-making. (Dhungana et al., 2025; Pepin et al., 2025) Especially, the communication of uncertainty has emerged as critical. Our results on enablers of successful uncertainty communication are broadly consistent with the existing literature. (Hadley et al., 2025b; McCabe et al., 2021) The Ebola modelling examples described by one participant illustrate how models from prestigious institutions, interpreted by decision-makers without sufficient technical literacy, can generate unwarranted confidence in projections that are later shown to be erroneous. This connects to concerns raised in the literature about the challenges non-technical audiences face in understanding modelling outputs (Dhungana et al., 2025; Pepin et al., 2025) and the risks of scientific authority being decoupled from model quality.

The heightened risks to modelling-informed decision-making in emergency settings is a particularly important finding. The time pressure of outbreak response limits the ability to triangulate modelling outputs with other evidence types, meaning that models may carry exceptional singular weight in shaping urgent decisions. Pre-established networks and institutionalized knowledge translation structures emerged from our data as critical safeguards: by enabling rapid, trust-based engagement between modellers and decision-makers, they reduce the risk of both model misspecification and communication failure under time constraints.

### A Three-Level Framework of Modelling-to-Policy Pathways

This study developed a three-level framework of factors influencing perceived successful modelling-evidence use starting from a systemic level and describes pathways and proposes mechanisms that link these factors between levels. Our analysis found these pathways to be structured around three interdependent domains: policy relevance, model quality, and communication and interaction (Figure 1). This framework is broadly consistent with existing conceptual work on facilitators of research contribution to policymaking that similarly identifies research quality and relevance (Cash et al., 2003), as well as mutual understanding and relationships and trust between knowledge producers and users as key enablers of evidence use. (Oliver et al., 2014; Sarkki et al., 2015) The risks to modelling-informed decision-making however emerged more prominently in our data, where we outline key factors and enabling inputs to prevent this. The factors we found facilitating modelling evidence use resonate with what has been described in the modelling-specific literature, such as the importance of high data quality, co-created and context-specific models, technical literacy of decision-makers and knowledge brokering capacities, stakeholder input and bidirectional communication (Figure 2). (Crouch et al., 2025; Hadley et al., 2025a; Moran et al., 2025; Owek et al., 2024; Rao et al., 2025) Our study contributes to this literature by spanning diverse geographic and institutional contexts, and by mapping the systemic enablers and mechanisms that give rise to the enabling conditions underpinning the domains of model quality, policy relevance and communication and interaction. It further goes beyond emergency decision-making – other than most of the modelling-to-policy literature that focusses on the COVID-19 pandemic context – and includes perspectives on routine decision-making as well.

Our findings further extend the typology of modelling-to-policy systems developed by Hadley et al. (Hadley et al., 2025a), which mapped configurations of how modelling teams and decision-making bodies relate to one another. This includes the number of and collaboration between modelling teams that inform national decision-makers, and their level of government interaction ranging from medium to embedded, i.e. modellers being employed by decision-making bodies. While Hadley et al.’s typology provides a valuable classification of system types, it does not explicitly account for settings in which modellers operate in geographically distinct locations from the decision-makers they inform – a configuration that emerged prominently in our data, particularly in low- and middle-income country contexts where international modelling teams frequently support national decision-making. Also, Hadley’s typology does not express a preference for any particular system type. (Hadley et al., 2025a) Our findings suggest, however, that geographic and institutional distance carries compounded risks around model misspecification and communication failure that locally embedded modellers are better placed to mitigate. Local and embedded modellers can draw on contextual knowledge, established trust, and direct iterative relationships with decision-makers that our data identified as key enablers across all three domains of our framework, and that can mitigate risks arising from inadequately specified models or communication failures through more contextualized outputs and direct, iterative communication. These advantages of local capacity and embeddedness have been argued for elsewhere in the literature (Aguas et al., 2020; Aheto et al., 2026; Moran et al., 2025; Oliwa et al., 2025; Owek et al., 2024; Thumbi et al., 2024), and our framework proposes a structured account of the mechanisms through which they operate.

A notable tension emerging from our findings is that institutionalized knowledge translation structures – while identified as systemic enablers – carry their own risks. By channelling access to decision-makers through established structures, they may inadvertently exclude modelling approaches and voices that sit outside them, narrowing the evidence base and potentially limiting how comprehensively policy questions are addressed. This resonates with Asatani et al.’s (Asatani et al., 2026) observation that when institutionalization takes place before the field has reached shared scientific ground, distinct legitimacy concerns arise about whose knowledge reaches decision-makers and under what conditions. Ensuring diversity in the composition of such structures – such as advisory boards – is therefore critical, yet particularly challenging when they are constituted ad hoc under time-constrained emergency conditions (Behdinan et al., 2018; Gopinathan et al., 2018). Institutionalized structures should therefore be understood not as endpoints but as ongoing achievements requiring critical scrutiny – designed with sufficient openness to accommodate methodological diversity and to remain responsive to evolving evidence needs. A distinctive contribution of our framework is that it represents, to our knowledge, the first multi-level account of modelling-to-policy pathways proposing mechanisms, developed from empirical qualitative data spanning diverse geographic and institutional contexts. Existing frameworks relevant to this space fall short in several respects. First attempts to map pathways specifically for modelling evidence have set out how scenario modelling can support decision-making (Androutsopoulou & Charalabidis, 2018), but this work is not empirically grounded and does not identify the influential factors or preconditions that shape whether modelling evidence is actually used. A separate body of literature has examined facilitators and barriers to infectious disease modelling evidence use in public health decision-making (Hadley et al., 2025a; Oliwa et al., 2025; Owek et al., 2024; Rao et al., 2025), but this work is largely confined to emergency and outbreak contexts and, while identifying relevant factors also on a systemic level, does not link them in the logical chains needed to understand how they lead to use and utility. By mapping the pathways from systemic enablers through enabling conditions to use factors, our framework offers a theory of change that can serve two applied purposes. First, it can guide the *evaluation* of existing modelling-to-policy systems, providing a structured basis for assessing where in the proposed logic chain failures occur and why. Second, it can inform the *design and set-up* of new modelling-to-policy systems, identifying systemic investments that can enable successful modelling-to-policy pathways. Although the proposed mechanisms linking systemic enablers, enabling conditions, and use factors represent hypotheses derived from our data, and require further testing through empirical application, this positions the framework as a practical tool for health system planners, evaluators, funders, and modelling communities seeking to build or strengthen modelling-to-policy infrastructure. Its explicit structure of proposed causal pathways makes it amenable to testing across diverse decision-making contexts, through which additional systemic enablers and pathways may be identified, and existing ones refined – progressively contributing to a more comprehensive theory of change for modelling-to-policy systems.

### Limitations

Our study is subject to several limitations: First, while we identified a range of systemic enablers, our findings should not be interpreted as a comprehensive inventory. Given the breadth and complexity of the systems under study, additional enablers likely exist beyond those captured in our data, and future research may uncover further systemic enablers not represented here. Second, our analysis yielded no systemic enablers specific to flexible or adaptable models. As adaptability was found important by others (Crouch et al., 2025), these enablers warrant further investigation. Third, the study included a limited number of knowledge users, and politicians were not represented among participants. Participating knowledge users were predominantly from high-income countries, thus dominating the knowledge user perspective in this study. This may have introduced blind spots regarding the priorities and experiences of knowledge users in lower-income settings. Fourth, although inter-coder agreement was high for code completeness, most of the coding was carried out by a single researcher. This concentration of analytical work in one individual may have introduced subjectivity, despite the checks in place. Fifth, no participants from South America were included in the study, though all other continents were represented.

## Conclusion

This study offers a structured account of what constitutes successful modelling-to-policy pathways and the systemic conditions that enable them, drawing on perspectives from diverse geographic and institutional contexts. The resulting framework moves beyond descriptive accounts of barriers and facilitators to propose a mechanistic, multi-level theory of change that can guide both the evaluation of existing systems and the design of new ones.

Several important directions for future research emerge from this study. Our framework proposes a causal account of how systemic enablers give rise to the conditions underpinning successful modelling-to-policy pathways. But it does not address whether the identified success factors and enabling conditions can be brought about through alternative causal chains, nor whether some are more important than others in particular decision-making contexts. The relative importance of individual framework components is likely to vary across settings. Future research should therefore apply the framework empirically through in-depth case studies spanning diverse policy and decision-making contexts, with the dual aim of testing its applicability and identifying the contextual factors that shape which causal pathways and factors are most important. Such work could ultimately support the development of a refined typology of modelling-to-policy systems, mapping not only structural configurations, but the causal dynamics and context-specific conditions that determine how and when modelling evidence successfully informs policy.

## Data Availability

All data produced in the present study are available upon reasonable request to the authors.

## DATA AVAILABILITY

The datasets generated and analysed during the current study are available from the corresponding author on reasonable request.

## ACKNOWLEDGEMENTS

The authors gratefully acknowledge the contributions of all study participants, whose time and expertise made this research possible.

## AUTHOR CONTRIBUTIONS

This authors’ contributions statement follows the Contributor Role Taxonomy (CRediT) where applicable. (National Information Standards Organization (NISO), 2022)

**Conceptualization:** KR, ENI, SF, JF, JH

**Data curation:** KR, ENI

**Formal analysis:** KR, ENI

**Funding acquisition:** KR, ENI, SF, JF, JH

**Investigation:** KR, ENI

**Methodology:** KR, JH

**Project administration:** KR, ENI, JF

**Supervision:** SF, JF, JH

**Validation:** HTF, KO

**Visualization:** KR, ENI

**Writing – original draft:** KR

**Writing – review & editing**: ENI, HTF, KO, SF, JF

