## Supplementary Files for "Developing an Evaluation Framework for Infectious Disease Modelling-to-Policy Pathways: A Qualitative Study Across Five Continents"

***Table 2****:* ***Level 1:*** ***Factors directly influencing modelling evidence use within the respective domains.*** *Each factor is categorized within a domain; some fall under multiple domains as is stated in the “Intersections” column.*

| Domain | Use Factor | Explanation | Intersections |
| --- | --- | --- | --- |
| Policy relevance | Policy questions amenable to modelling | Modelling outputs are most relevant when they address policy questions that can realistically be answered through modelling approaches. |  |
|  | Context-specific models | Models reflect the epidemiological, demographic, and health system context they aim to inform. | Model quality |
|  | Timely outputs | Modelling results are delivered within the timeframes required for policy decisions. |  |
|  | Co-created modelling | Integrating users’ and relevant stakeholders’ perspectives in the development of models increases the likelihood that outputs meet policy needs and get integrated in decision-making. | Model quality  Communication and interaction |
|  | Adaptable and reusable models | Models are designed with flexibility to allow staff in decision-making bodies to adapt them to evolving policy needs without commissioning entirely new models. The ability to conduct their own analyses increases their ownership of the results. |  |
| Model quality | Adequate and justifiable assumptions | Model assumptions are adequate and justifiable. This requires expert disease and contextual knowledge and multidisciplinary perspectives. |  |
|  | Peer-reviewed models | Modelling outputs are reviewed formally or informally within the scientific community before informing policy. |  |
|  | Methodological diversity | Multiple modelling approaches are applied to the same policy question. This reduces structural uncertainty and increases utility for decision-makers. (Structural uncertainty is stemming from uncertainty in choices about the model structure.) | Policy relevance |
|  | Holistic modelling | We term models that integrate considerations from fields beyond health as “holistic”. This includes integrating economic impacts, mechanisms from behavioural sciences, and stakeholder knowledge. This multidisciplinary grounding, enriched by the perspectives of those with lived experience of policy impacts, allows models to capture interdependencies, feedback loops, emergent dynamics and potential impacts that characterise the policy problems the models aim to address. By explicitly acknowledging, integrating, and analysing real-world complexity, the modelling becomes more relevant for policy. | Policy relevance |
|  | Equity considerations | Questions of equity should be considered in the modelling processes, and modelling choices should be made such that these questions can be answered. This reduces the risk of misinforming policy decisions through the lack of consideration of sub-populations or minority groups. | Policy relevance |
| Communication and interaction | Adequate communication of uncertainty | Model assumptions, uncertainties, nuances, and limitations are openly communicated and in ways that are understandable to decision-makers. The adequate format of communication depends on the audience, their time constraints, and level of technical knowledge. | Policy relevance |
|  | Iterative and bi-directional interaction | Continuous dialogue between modellers and decision-makers allows for feedback and refinement of modelling questions and outputs. This can prevent misinformed decision-making and enhance mutual understanding and facilitate the co-creation of outputs. | Policy relevance |
|  | Sustained engagement | Sustained interaction between modellers and decision-makers build long-term relationships, trust and facilitate future engagement. Long-term relationships between modellers and decision-makers in turn support ongoing collaboration through improved communication channels and interaction. |  |
|  | Direct communication channels | Direct interaction minimizes loss of nuance and facilitates clarification of modelling results. This can prevent misinformed decision-making and enhance mutual understanding. It can also improve decision-makers’ ownership and buy-in. |  |

Table 3 summarises our findings on how perceived successful modelling evidence use can be enabled, outlining the influential factors in a three-level structure. In the first column, we identify various enablers on a system-level. These systemic enablers facilitate conditions (second column) that in turn enable use factors (third column) (Figure 1). The fourth column describes the proposed causal mechanisms of how the systemic enablers and enabling conditions lead to the use factors. The presented pathways and factors comprehensively summarize what resulted from the analysis of our data. However, there might be other systemic inputs that could lead to the enabling conditions and use factors, that were not covered in our data.

**Table 3: Three-level framework outlining systemic enablers, enabling conditions, enabled use factors and the proposed causal mechanisms linking them.**

| Level 3: Systemic Enabler | Level 2: Enabling Condition | Level 1: Enabled Use Factor(s) | Proposed Mechanism |
| --- | --- | --- | --- |
| Local modelling capacity | Contextual knowledge | - Context-specific models - Adequate assumptions | Local modelling teams possess the contextual knowledge – spanning disease dynamics, health systems including data infrastructure, and governance environments – necessary to develop context-specific models grounded in adequate assumptions. |
|  | Trust between modellers and decision-makers | - Direct communication channels - Iterative and bi-directional interaction - Adequate assumptions - Timely outputs - Sustained engagement | Local modellers may enjoy greater trust among governmental actors, facilitating data sharing, more adequate assumptions, timely outputs, and sustained engagement over time. However, this trust is contingent on local modellers demonstrating sufficient technical expertise, as decision-makers' confidence in their capabilities is a prerequisite for a productive working relationship. In this respect, internationally renowned institutions may carry greater inherent credibility, which can discourage decision-makers from engaging with local modellers altogether.  Trust between modellers and decision-makers can motivate more direct and iterative interaction. Local modelling capacities can thereby also serve as a bridge to international modelling teams, supporting the communication and engagement with decision-makers that international modellers may find difficult to establish independently. |
| High quality data and well-functioning data infrastructure | Data availability and quality | - Adequate assumptions - Context-specific models | Reliable data enable more accurate parameterization and modelling assumptions relevant to the specific context. |
|  | Granular analyses | - Equity considerations | Comprehensive datasets with granular information on specific population groups - including minorities – enable targeted analyses that support equity considerations in modelling outputs. |
| Knowledge brokering capacity | Mutual understanding between modellers and decision-makers | - Adequate communication of uncertainty - Policy questions amenable to modelling | Knowledge brokers or intermediaries with insight into both the technical and policy domains can bridge gaps in mutual understanding between modellers and decision-makers. This reduces the risk of misinterpretation and misinformation, while increasing the likelihood that decision-makers draw on modelling evidence for questions that are amenable to modelling. |
|  | Linkage to external modellers | - Iterative and bi-directional interaction - Sustained engagement | Knowledge brokers with a higher proximity to decision-makers can highlight relevant modelling evidence from external modellers, provide the latter with access to decision-makers, and facilitate their iterative, bi-directional interaction and sustained engagement that external modellers would otherwise struggle to establish independently. |
| Embedded modelling capacity / smaller country contexts | Low-threshold communication channels  Trust between modellers and decision-makers  Incentivized modellers | - Iterative and bi-directional interaction - Direct communication channels - Policy questions amenable to modelling - Co-created modelling - Timely outputs - Sustained engagement | Modelling capacities embedded within decision-making institutions facilitate trust between modellers and decision-makers, supported by low-threshold communication channels that enable direct, iterative, and bi-directional interaction. Regular communication facilitates the refinement of modelling questions, clarification of uncertainties, and mutual understanding, while trusted relationships support sustained, long-term engagement. In smaller countries, the more limited number of stakeholders involved in modelling-to-decision-making processes can enable comparable levels of relational proximity even without in-house modelling capacities. Additionally, modellers embedded within – and funded by – decision-making institutions may have stronger incentives to prioritise policy-relevant outputs and deliver them in a timely manner. |
| Institutionalized/formal knowledge translation structures (e.g. advisory boards, policy dialogues) | Institutionalized communication channels  Trust between modellers and decision-makers  Resilient communication channels | - Iterative and bi-directional interaction - Direct communication channels - Co-created modelling - Sustained engagement | Formal interaction and communication structures foster relationship-building and trust through repeated engagement. This can enable iterative and bi-directional interaction, allowing for co-creation of models, while also opening more direct and informal channels of communication. Trust also supports sustained engagement. This is particularly relevant for external, e.g. academic, modellers who are not embedded within decision-making institutions.  Formal, institutionalized structures can increase the resilience of interaction and communication channels between modellers and decision-makers and their collaborations to changes in government and thus enable sustained, long-term engagement. |
| Training for modellers on communication and policy needs  Modelling literacy among decision-makers | Mutual understanding between modellers and decision-makers | - Adequate communication of uncertainty - Policy questions amenable to modelling | A better understanding of policy needs and communication skills among modellers allow for adequate communication of uncertainty that prevents misinformation. Modelling literacy among decision-makers enables them to assess what questions modelling can and cannot answer, to interpret outputs critically and to appreciate caveats and uncertainty that prevents the misinterpretation of outputs. |
| Established networks among modelling teams, experts from other disciplines and policy actors (e.g. consortia, online collaboration platforms) | Collaboration between different modelling teams | - Peer-reviewed models - Adequate assumptions - Methodological diversity | Established networks facilitate joint modelling efforts and informal peer review. They enable methodological diversity and appraisal of structural uncertainty when multiple teams apply different modelling approaches to the same policy question. |
|  | Expert and stakeholder input | - Holistic modelling - Equity considerations - Adequate assumptions | Connections to experts from other disciplines – such as behavioural scientists, economists, and sociologists – as well as engagement with relevant stakeholders and practitioners, facilitate the integration of diverse perspectives into model development. This broadens the conceptual scope of modelling efforts, counteracts disciplinary tunnel vision, and enables more holistic, systems-oriented representations of real-world complexity. Subject-matter expertise improves the quality of modelling assumptions, while stakeholder engagement helps ensure that models capture impacts across diverse population groups. |
|  | Access to modelling surge capacity | - Timely outputs | Established networks among stakeholders involved in modelling-to-decision-making processes – including decision-makers, modellers, and knowledge brokers – can facilitate access to existing models and reduce redundancy more broadly, while also enabling the rapid scale-up of modelling capacities in emergency situations. This increases the likelihood of timely outputs in fast-moving contexts such as infectious disease outbreaks. Such networks may operate at both national and international levels. |
|  | Trust between modellers and decision-makers | - Sustained engagement - Timely outputs | Established networks foster relationship-building among actors through continuous interaction, building trust that supports sustained engagement over time.  Trusted relationships facilitate data sharing, also internationally, enabling timely outputs. |
| Diversity within modelling teams | Diversity of perspectives | - Holistic modelling - Equity considerations | Diversity within modelling teams – across gender, ethnicity, and geographic and cultural backgrounds – broadens the range of perspectives brought to bear in model development, increasing the likelihood of producing outputs that are holistic and equitable. |
| Possibilities for modellers to publish | Incentivized modellers | - Timely outputs - Sustained engagement | For academic modelling teams in particular, agreements with decision-makers on publication rights – for instance where decision-makers hold ownership of the underlying data – can serve as an important incentive to engage with policy-relevant questions, produce timely outputs and continuously engage with decision-makers. |

### Topic guide: Knowledge producers

**Theme 1: Modelling – Policy Interaction**

Have you produced modelling evidence for policy or other decision-makers in the past?

- What policy questions were that?
  - In which context was that? Can you give examples?
  - Which policy or decision-making bodies did you inform?
    - *NGO, regional, national, international*
    - *Differences?*
- Why did you engage in modelling for decision-making?
  - *Motivation? Capabilities, opportunities?*
  - Who initiated the contact? Why? How?
    - *Platform? Pre-existing relations?*
    - *Commissioned? Did you send your results?*
    - *Time point of start of interaction? Co-creation?*
- Who funded your modelling efforts then?
  - *Policymakers? Continued? Changed?*

How did you interact with policy- or decision-makers in the past? Can you give examples?

- Differences? Why? What did work well?
- To whom did you talk to? Directly to decision-makers?
  - *Decision-making hierarchy dependent?*
  - *Knowledge brokers?*
    - *Influencing comprehensibility?*
- How often?
  - *How many teams? Frequency? Inhouse / external? International / national? Channels? Timepoint start of engagement?*
- Did you interact or collaborate with other modellers / modelling teams? Why? What for?
  - *How? Pre-established networks? Platforms?*
- Have you received feedback by policymakers? Can you give examples?
  - When? How often? What about?
  - Is this helpful for your work? How and why?
  - Increased relevance?

How did you communicate uncertainty?

- Differences according to interaction structure?
- Do you feel information was lost or misunderstood sometimes?
  - Why? In which contexts?
  - *Decision-making hierarchy, knowledge brokers, no direct interaction*

Did you feel some interactions were more successful/useful? Why?

- *Other factors influenced by infrastructure and knowledge translation*
- What makes the interaction successful in your opinion?
  - *Different motivations?*

**Theme 2: Modelling**

How did you try to ensure that your results are relevant for policymaking?

- When? Depending on what?
  - *Modelling infrastructure, commissioning, motivation, policy question/context?*
- How did you assess uncertainty in the model and the results?
  - *Data?*
- What else did you consider to ensure relevance of your results?
  - *Quality, co-creation, early on communication, commissioning, timeliness, collaboration with stakeholders, collaboration with other disciplines*

Did you need to make tradeoffs between model quality and other factors? Can you give examples?

- *E.g. timeliness?*
- Differences according to policy question/context? *Outbreak vs. routine?*
- How did you try to improve model quality while still producing relevant results?

Reflecting on what we have discussed so far, what would you say is the general purpose of modelling for decision-making?

- What would you define as success?
- Can you give examples when that was achieved?
- What hindered this in your experience?
  - *Would you do anything differently? What? Why?*

**Theme 3: Equity and Outlook**

Have you ever considered the impact in different communities or minority populations in your models to inform decision-making?

- Can you give an example?
- What were the consequences of not considering equity aspects in your experience?

What are your future plans in terms of engaging in providing modelling evidence to inform decision-making?

- Why?
- What would make future engagement more likely / easy?
- *Maintenance of structures / relations / communication channels?*
- *Maintenance of collaboration with other modellers / stakeholders?*

Is there anything else you would like to add?

### Topic guide: Knowledge users

Since this is a semi-structured interview, a selection of the following questions will be used to generate discussion.

**Theme 1: Modelling – Policy Interactions**

Have you referred to modelling for your work in the past?

- In which context was that? What decisions were these?
  - Can you give examples?
- Why did you draw on modelling evidence for your decision-making?
  - *Motivation, trust in modelling evidence?*
  - *Awareness about modelling infrastructure and perceived value of modelling?*
  - How did you receive it? Can you give examples?
    - Did you commission it or was it sent to you?
      - *Influence of commissioning on relevance?*
      - *Timepoint of start of interaction with modeler?*
    - Who funded the modelling efforts? Why?
      - *Continued funding? Changes in funding?*
- How did you interact with modellers when you drew on modelling evidence? Examples?
  - *Directly?*
    - *Decision-making hierarchy dependent?*
  - *Knowledge brokers?*
    - *Re-translation?*
  - *How many teams? Frequency? Inhouse / external? International / national? Channels? Feedback loops? Timepoint start of engagement?*
  - Differences? Why?
- Did you feel some interactions/structures were more successful/useful? Why?
  - *Factors influenced by infrastructure*
  - *Knowledge brokers influencing comprehensibility?*
  - *Re-translation: Risk of losing information?*
- How was modelling evidence communicated to you?
  - *Channels? Comprehensibility?*
  - How was uncertainty communicated?
    - Differences according to interaction structure?
  - Do you feel information was lost or misunderstood sometimes?
    - Why? In which contexts?
    - *Decision-making hierarchy, knowledge brokers, no direct interaction*

**Theme 2: Modelling Evidence Use in Decision-making**

Did modelling evidence help you in your decisions in the past? If so, how? Can you give examples?

- *Use and utility*
- *Did you take decisions based on modelling evidence?*
- Has modelling evidence changed your opinion on policy problems in the past?
  - How?
  - *Opinion change?*
- *Symbolic, enlightenment, conceptual, instrumental, re-contextualization?*

Why do you think modelling evidence was useful? Why not?

- *Added value?*
- Were there differences in how helpful the evidence was? Why? Which contexts? Examples?
  - *Timeliness?*
  - *Policy question? Outbreak vs. routine?*
  - *Availability of other information of higher quality?*
  - ***Modelling-policy interaction (structures, low to embedded)***
  - *Uncertainty? Quality?*
  - *Uncertainty communication?*
  - *Relevance? Timeliness? Co-creation?*
  - *Context-specificity? Collaboration with different stakeholders?*
- What are situations where it was less useful? Why?
- How did it compare to other evidence?
- *How balance modelling evidence with other sources of information / types of evidence?*
- *Decision-making in the absence of modelling evidence?*

What decisions did drawing on modelling evidence lead to? Did it lead to different decisions?

- Can you give examples?
- Why different decisions?
  - *What makes successful outcomes more likely?*
- Sometimes no decision at all? Why?
- Did it sometimes lead to unjustified decisions? Why?
  - *Over-persuasion / misinformation?*
  - *Uncertainty communication?*

**Theme 3: Equity and Outlook**

Have you ever considered the impact in different communities or minority populations in your decision-making? Do you consider potential unintended consequences of decisions?

- Have you used modelling evidence for that?
- Can you give an example?
- What were the consequences of not considering equity aspects in your experience?

What are your future plans in terms of engaging in using modelling evidence to inform decision-making?

- Why?
- What would make a future engagement more likely / easy?
- *Maintaining funding / structures / relationships?*

Is there anything else you would like to add?
